# Severe acute myositis and myocarditis upon initiation of six-weekly Pembrolizumab post-COVID-19 mRNA vaccination

**DOI:** 10.1101/2023.11.24.23296021

**Authors:** Robert A. Watson, Weiyu Ye, Chelsea A. Taylor, Elsita Jungkurth, Rosalin Cooper, Orion Tong, Tim James, Brian Shine, Monika Hofer, Damian Jenkins, Robert Pell, Eleni Ieremia, Stephanie Jones, David Maldonado-Perez, Ian S.D. Roberts, Nicholas Coupe, Mark R. Middleton, Miranda J. Payne, Benjamin P. Fairfax

**Affiliations:** MRC Weatherall Institute of Molecular Medicine, University of Oxford, Oxford, UK; Department of Oncology, University of Oxford, Oxford, UK; Cancer and Haematology Centre, Oxford University Hospitals NHS Foundation Trust, Oxford, UK; Department of Clinical Biochemistry, Oxford University Hospitals NHS Foundation Trust, Oxford, UK; Department of Neuro Pathology, Oxford University Hospitals NHS Foundation Trust, Oxford, UK; Department of Clinical Neurology, Oxford University Hospitals NHS Foundation Trust, Oxford, UK; Department of Cellular Pathology, Oxford University Hospitals NHS Foundation Trust, Oxford, UK; Oxford Centre for Histopathological Research, Oxford University Hospitals NHS Foundation Trust, Oxford, UK

## Abstract

We describe three cases of critical acute myositis with myocarditis occurring within 22 days of each other at a single institution, all within one month of receiving the initial cycle of the anti-PD-1 drug Pembrolizumab. Analysis of T cell receptor repertoires from peripheral blood and tissues revealed a high degree of clonal expansion and public clones between cases, with several T cell clones expanded within the skeletal muscle putatively recognising viral epitopes. All patients had recently received a COVID-19 mRNA booster vaccine prior to treatment and were positive for SARS-CoV2 Spike antibody. In conclusion, we report a series of unusually severe myositis and myocarditis following PD-1 blockade and the COVID-19 mRNA vaccination.

## INTRODUCTION

Immune checkpoint blockade (ICB) with anti-PD-1 monoclonal antibody is approved for treatment of melanoma in adjuvant and palliative settings^1,2^. These treatments can elicit immune-related adverse events (irAEs)^3,4^ including myositis and myocarditis - although the occurrence of these particular toxicities is very rare, being observed in <1% of cases with fatalities reported in <0.01% of recipients of anti-PD-1 alone^5,6^.

Infection with SARS-CoV2 can lead to both myositis and myocarditis^7^. Whilst the underlying mechanisms remain undetermined, indirect virally-triggered autoimmune reaction or direct epitope cross-reactivity are posited^8^. There is also an association between SARS-CoV2 vaccination with mRNA vaccines and myocarditis. This is rare, appears to be driven by a younger patient population and the underlying mechanisms are currently unclear^9–12^.

## CASE REPORTS

### Patient 1

A patient in their early 70s with pre-treatment ECOG performance status (PS) 0 presented with dizziness and dyspnoea 28 days post-initiation of Pembrolizumab (six week infusion, 400 mg) for adjuvant treatment of resected Stage IIIC melanoma. Medical history consisted of atrial fibrillation and type 2 diabetes. They attended a nearby hospital five days prior for non-specific chest pain, treated with analgesia. On arrival, they reported lethargy, weakness and inability to support his head, with associated bulbar symptoms. They had no chest pain or ischaemic ECG changes, but serum creatine kinase (CK) and troponin were markedly elevated, (CK: 2,236 U/L, range 30-200 U/L; Troponin: 267 ng/l, range 0-34 ng/l; Figures 1a, 1b and Table 1) as were liver enzymes (alanine aminotransferase, ALT, 394 U/L, range 10-45 U/L; alkaline phosphatase, ALP, 1,044 U/L, range 30-130 U/L) and bilirubin (68 umol/L, range 0-21 umol/L). Their condition acutely worsened, developing type II respiratory failure with reduced consciousness, precipitating transfer to critical care with vasopressor support and ventilation. Initial management was with intravenous methylprednisolone (2mg/kg) for five days but upon further deterioration, intravenous immunoglobulin (IVIg) was given for a further five days. An electromyogram demonstrated a necrotic myopathic process involving proximal limbs with background neuropathy. Autoantibody screening revealed positivity for anti-acetylcholine receptor antibodies (AChR). A muscle biopsy showed multifocal clusters of necrotic fibres, consistent with an ICB-associated myositis^13^ (Figure 1c). The final diagnosis was Pembrolizumab-associated myositis, myocarditis and myasthaenia gravis with hepatitis. Pyridostigmine was commenced, with little initial benefit, followed by plasma exchange (commenced 3 weeks following IVIg). A slow but sustained clinical improvement ensued and they were discharged to a rehabilitation unit. They received a booster vaccination (BNT162b2) 28 days prior to Pembrolizumab.

**Figure 1.**
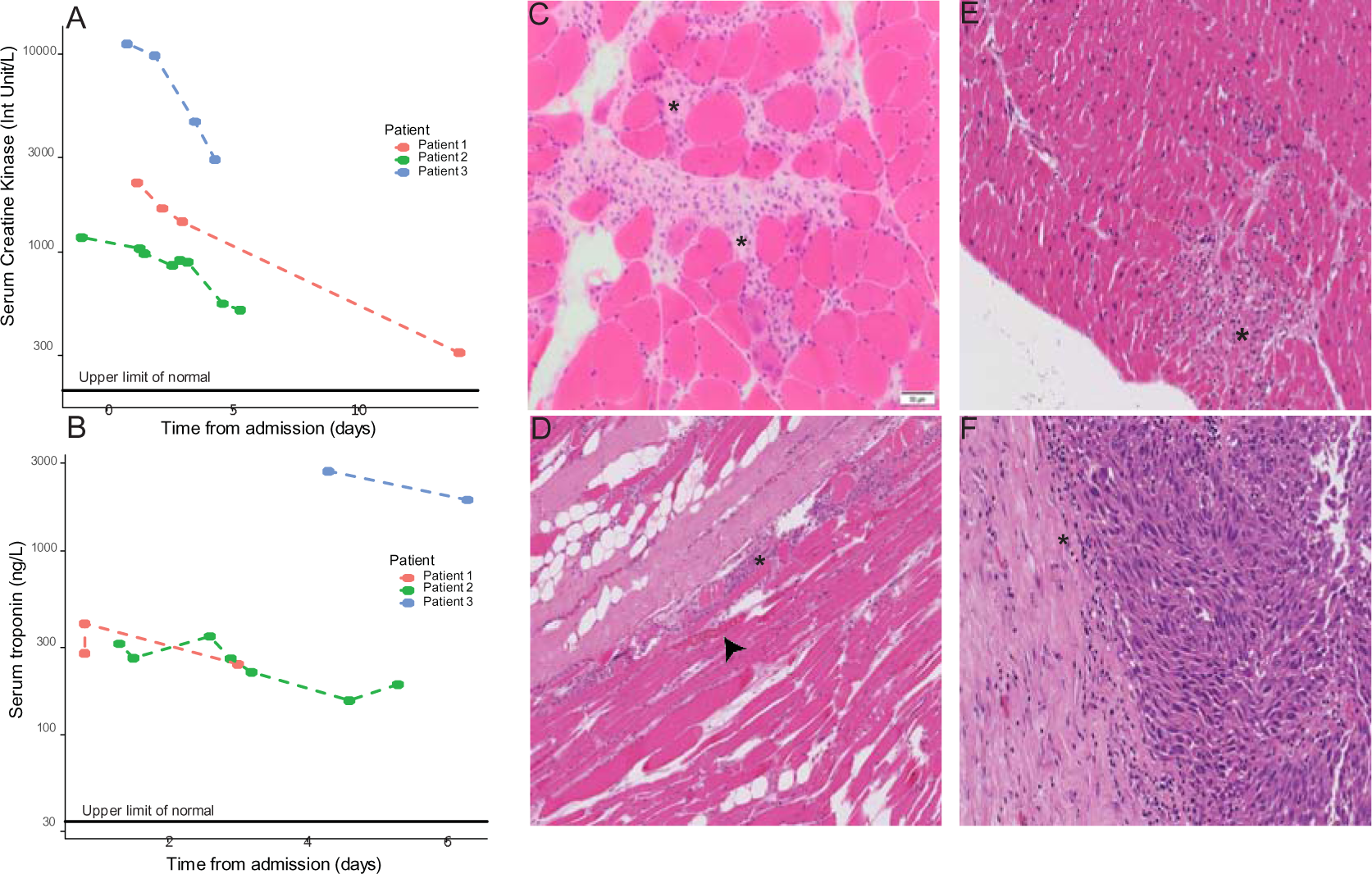
(A) Serum creatine kinase (CK) by day post-admission for each of the three patients. (B) as per (A) but for troponin. (C) Haematoxylin and Eosin (H&E) slide of fresh skeletal muscle biopsy taken from patient 1. Widespread leukocyte infiltration can be seen (e.g. at asterixes). (D) H&E stained slide of post-mortem (PM) skeletal muscle sample taken from patient 3. The asterix indicates infiltrating lymphocytes with the arrowhead denoting myocyte necrosis. (E) H&E slide of post-mortem cardiac muscle from patient 3, demonstrating widespread leukocyte infiltration. (F) as per (E) but for tumour deposit taken from the small bowel serosa.

**Table 1.**
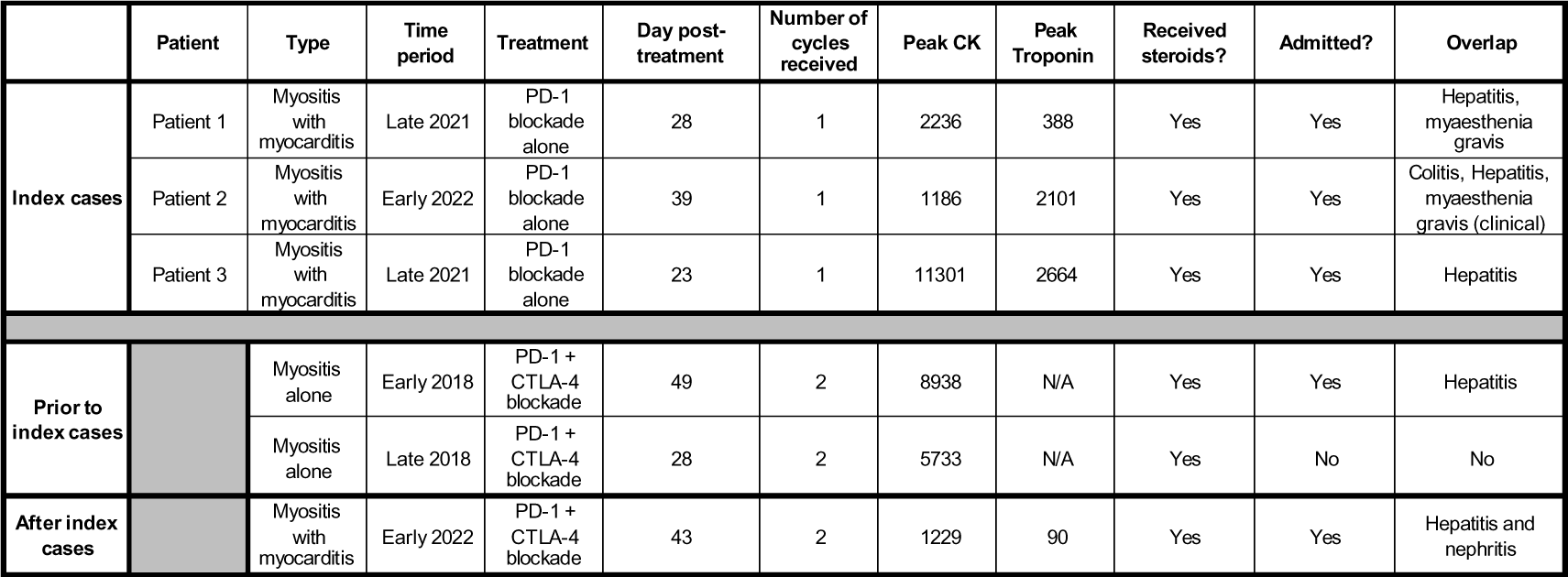
Summary of cases of myositis or myocarditis, comparing index cases (top section) to those found within a cohort of 237 patients from end 2015-end 2021 (‘prior to index cases’, bottom section) and a further case identified subsequent to the index cases in early 2022.

### Patient 2

A patient in their 80s with pre-treatment PS 0 and no relevant medical history was admitted 39 days post-initiation of Pembrolizumab (400mg) for stage IV melanoma. They developed diarrhoea and severe myalgia 11 days after treatment, attending a local hospital where oral prednisolone (60mg) was commenced. Subsequently, the diarrhoea resolved whereas the myalgia worsened and on day 23 she was re-admitted having collapsed with loss of consciousness. They were in complete heart block but without ECG features of acute ischaemia, however serum troponin was elevated (1,124 ng/L). Coronary angiography demonstrated a possible ruptured left anterior descending artery plaque and she was managed with percutaneous stent placement and a permanent pacemaker. Despite this, blood tests six days post-discharge showed further elevation of serum troponin (2,101 ng/L) and elevated CK (1,186 U/L) (Figure 1a, 1b and Table 1) which precipitated admission to our centre (day 39 post-treatment). ALT was also mildly elevated (216 U/L), ALP was normal. A clinical diagnosis of immunotherapy-associated myositis and myocarditis was made and intravenous methylprednisolone was given for three days, followed by oral prednisolone with slow clinical and biochemical improvement. However, ten weeks post-discharge they were readmitted with increasing fatigue, weakness, dysarthria, diplopia and ptosis. AChR antibodies were negative. A clinical diagnosis of Pembrolizumab-associated ocular myopathy and myositis was made, along with possible myasthenia; management was with oral corticosteroids, a course of IVIg and physiotherapy rehabilitation. They received a booster vaccination (BNT162b2) 74 days prior to Pembrolizumab.

### Patient 3

A patient in their early 80s with pre-treatment PS 0 was admitted with five days of reduced mobility, fatigue and myalgia (on movement but not palpation) 23 days post-initiation of Pembrolizumab (400mg) for stage IV melanoma. CK was markedly elevated (11,301 U/L) (Figure 1a, Table 1), as were liver enzymes (ALT 705 U/L, ALP 155 U/L). Troponin, measured on day 4 post-admission was 2,931 ng/L (Figure 1b, Table 1). Management was with intravenous methylprednisolone, switched to oral prednisolone after four days. Supplemental oxygen was started but weaned due to symptomatic improvement and decline in CK (Figure 1a) over four days, although bulbar symptoms and dysphagia then developed. An echocardiogram performed five days post-admission demonstrated normal bi-ventricular systolic function. Six days post-admission, they acutely deteriorated with type II respiratory failure and subsequent cardiac arrest. A post-mortem (PM) examination demonstrated multiple foci of inflammation and myocyte necrosis throughout the myocardium. Replacement fibrosis was absent, indicating a two-week timeframe, and there was minimal atheroma and no myocardial infarction. Skeletal muscle examination demonstrated multiple foci of inflammation and necrosis, similar to the sampled myocardium. Of note, a metastatic tumour deposit from the serosal surface of the small bowel displayed brisk lymphocyte infiltration (Figure 1d, 1e, 1f). They had received a booster vaccination (BNT162b2) 74 days prior to Pembrolizumab.

All three patients were negative for SARS-CoV2 anti-nucleocapsid IgG and positive for anti-spike IgG.

## METHODS

We performed an analysis of immune-related adverse events (irAEs) occurring within a cohort of patients receiving ICB (both anti-PD-1 and anti-PD-1/CTLA-4 combined at varying doses) for melanoma and renal cell carcinoma since 2015^3,14–16^. Patients provided written informed consent for the donation of tissue and access to clinical records to the Oxford Radcliffe Biobank (ethical approval 19/SC/0173, projects 16/A019, 18/A064, and 19/A114).

We analysed T cell receptor (TCR) repertoires from peripheral blood and tissue taken from the three patients reported here. PM tissue was obtained with informed consent from relatives and under institutional ethical approval (CUREC-1, R80630/RE001).

Blood collected into EDTA-coated tubes was separated using density centrifugation (Ficoll Paque) with plasma removed and ultra-centrifuged. Routine biochemistry tests were undertaken on thawed plasma samples using the Abbott Architect c16000; hs-troponin and COVID antibodies using an Abbott Architect i2000. Whole PBMCs and magnetically-sorted CD8^+^ T cells (MACS system, Miltenyi Biotec) were lysed in RLT Plus buffer supplemented with 40mM DTT, homogenised using QIAshredder columns prior to RNA and gDNA extraction using AllPrep DNA/RNA/miRNA Universal extraction kits (Qiagen). Fresh skeletal muscle was flash-frozen at −80°C before RNA was extracted using the RNeasy Plus Universal Mini Kit (Qiagen). RNA was extracted from paraffin-embedded PM tissue using the AllPrep FFPE kit (Qiagen). TCR repertoire libraries were constructed using the QIAseq Immune Repertoire library kit (Qiagen). Sequencing was performed on a MiSeq (Illumina) with pre-processing and alignment using the CLC genomics workbench (Qiagen). CDR3B chains were matched to epitopes using The Immune Epitope Database (IEDB) TCRMatch tool (http://tools.iedb.org/tcrmatch/) with the highest scoring epitope match being assigned to each CDR3B chain. All downstream bioinformatic and statistical analysis was performed in R (v4.0.5).

## RESULTS

### Comparison with cohort from this centre

We examined the incidence of myositis and myocarditis across a cohort of patients receiving ICB for melanoma and renal cell carcinoma from end 2015 - end 2021^3,14–16^ (n=135 who received combination programmed cell death protein 1 (PD-1) and cytotoxic T-lymphocyte-associated antigen 4 (CTLA-4) blockade, n=102 who received single-agent anti-PD-1). In line with published incidence^6^ only two of the 237 patients suffered biochemically-proven myositis and no myocarditis cases were recorded. Both patients had received two doses of *combined* PD-1/CTLA-4 blockade (cICB) (Table 1). After the occurrence of the three index cases which form this series, we identified a further individual in our cohort who developed clinical myositis with an asymptomatic troponin rise after his second cycle of cICB for metastatic renal cell cancer, 66 days post-booster (BNT162b2). This culminates in four cases of myositis with likely myocarditis within 28 days over the winter of 2021-2022, on a background of only two cases of myositis without myocarditis occurring within the same patient cohort over the preceding six years. In the three index cases we screened serum for the development of muscle specific immuno-reactivity with standard of care immuno-blots against cN-1A, MDA-5, Tif1-gamma, NXP-2, SAE-1, Mi-2a, Mi-2b, Ku, PM-Scl 100, PM-Scl-75, Jo-1, SRP, PL-7, PL-12, EJ, OJ and Ro52 - but there was no evidence of seropositivity, excluding development of antibodies towards common myositis antigens and suggestive of T cell mediated toxicity.

### Analysis of T cell repertoires

T Cell Receptor sequencing (TCRseq) of peripheral blood and tissue samples from all three patients (detailed in table S1) was performed, identifying a high degree of clonal sharing both within individuals (different tissue sites) and between individuals (Figures 2, S1 and S2). Clonally-expanded TCR present in the muscle biopsy from patient 1 were found similarly expanded within the peripheral blood, with a trend towards greater muscle expansion (Figure 2a). Notably, there was high clonal overlap between TCR found in the PM tumour specimen from patient 3 and the other sampled tissue sites (Figures 2b, 2c and S2). Of the 13 TCR found within the PM cardiac muscle of patient 3, 5 matched those found within the skeletal muscle biopsy of patient 1 (Figure 2c) and one was found in the peripheral blood of patient 2 (not shown).

**Figure 2.**
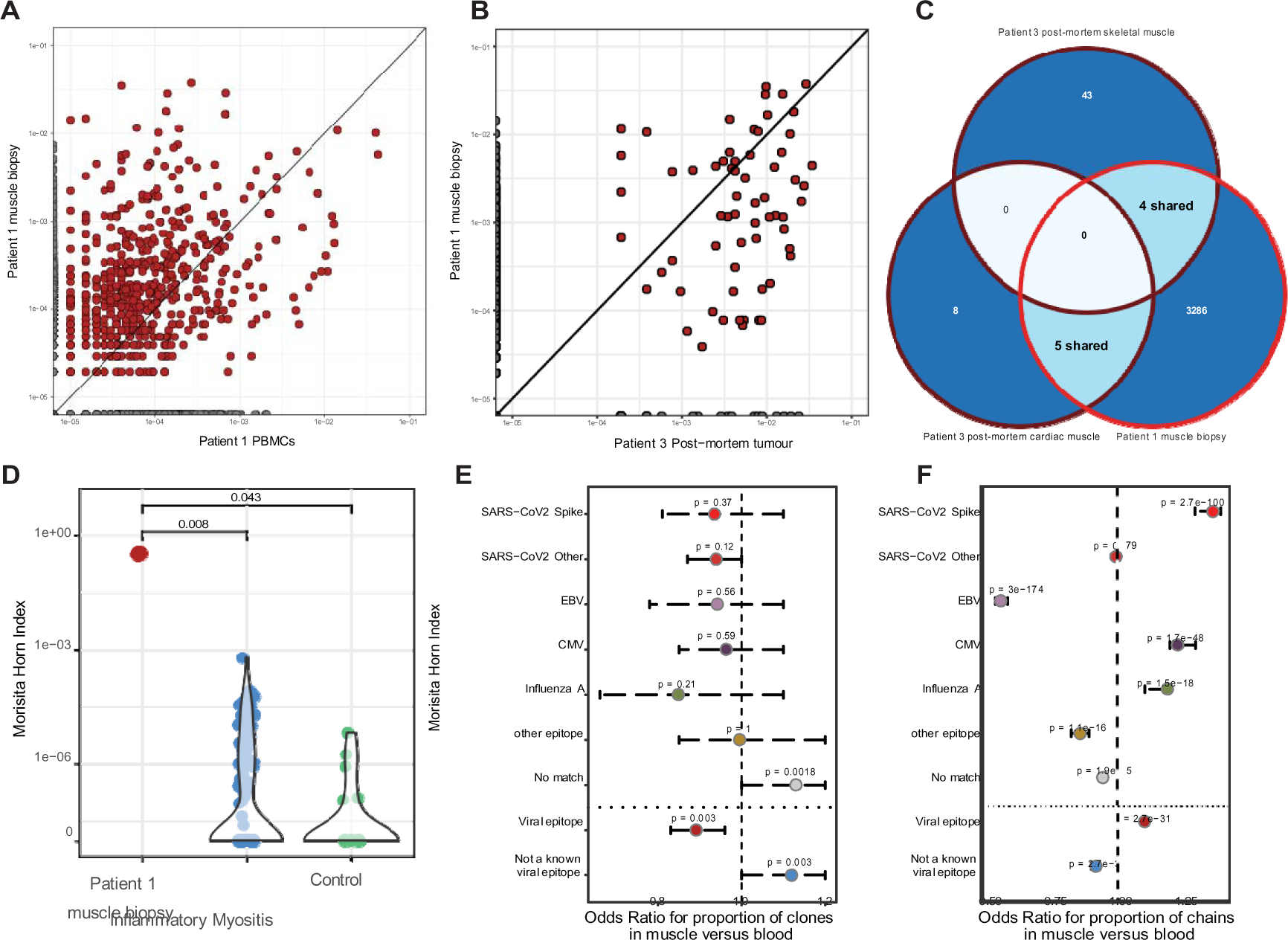
(A) Dot plot showing proportion of repertoire occupied by T cell clone in peripheral blood versus muscle biopsy from patient 1. (B) as for (A) but comparing the tumour from patient 3 with the muscle from patient 1. (C) a Venn diagram of the TCR overlap between cardiac and skeletal muscle of patient 3 and skeletal muscle of patient 1. (D) Morisita-Horn (MH) index for the repertoire overlap between the resected tumour from patient 3 and the skeletal muscle of patient 1, and samples taken from patients with IIM^18^. (E) Odds ratio for occurrence of epitope-specific clones (TCR) found in the muscle versus the peripheral blood of patient 1. (F) As per (E) but taking into account number of copies of each clone (thereby considering clonal expansion). Statistics are via the Kolgomorov Smirnov test (D) or Fisher’s exact test (E-F).

To quantify repertoire overlap, we applied the Morisita-Horn (MH) similarity index^17^. This confirmed high overlap between the TCR repertoire from the PM tumour sample from patient 3 and the fresh muscle biopsy from patient 1 (MH 0.328) (Figure 2d). This was significantly greater than the overlap between the PM tumour and TCR repertoires sequenced from muscle biopsies taken from controls and patients with idiopathic inflammatory myositis (IIM)^18^ (median MH for IIM samples 0, IQR 0-8.72e-7, p=0.008) (Figure 2d), further illustrated by plotting clonal sharing (Figure S3). We explored the overlap of the TCR repertoires from resected melanomas from eight patients in our original cohort (all resections pre-2020, Table S2). There was limited overlap and clonal sharing between these tumours and the muscle of patient 1, which was the same as their overlap with IIM samples (Figures S4, S5a). A similar finding was noted when comparing the TCR found within the cardiac tissue of patient 3 and either IIM muscle biopsies or previously resected tumours (Figures S5b, S5c).

Finally, we examined the nature of putative TCR epitopes from specimens taken from patient 1, with multiple shared clones matching a range of viral epitopes (Table S3). We tested for the enrichment of each clone within the muscle versus the peripheral blood, finding that although no significant differences in total unique clones recognising specific epitopes (Figure 2e), when taking into account clone size, TCR putatively recognising the SARS-CoV2 Spike protein were enriched within the muscle (OR 1.37, 95% CI: 1.30-1.40, p<0.0001), along with TCR reactive to viral epitopes in general (Figure 2f).

## DISCUSSION

We describe a cluster of three highly unusual cases presenting with clinically significant and life-threatening acute myositis with cardiac involvement within a three-week window, post-receipt of the first six-weekly dosage of Pembrolizumab. Pharmacovigilance studies of immunotherapy associated myocarditis show a higher incidence and increased severity in recipients of combination anti-CTLA-4 and anti-PD-1 immunotherapy, whilst approximately 25% of cases show evidence of myositis and 10% have features of myaesthenia gravis^19^. Occurrence of all three is referred to as the ‘3M syndrome’, has a much lower incidence than myocarditis alone, and is more severe^20^. Strikingly, all three described cases had characteristics of the ‘3M’ syndrome. None of the attending physicians had witnessed such rapid-onset and severe myositis with respiratory, cardiac and bulbar involvement post-ICB over many previous years of practice, underlining the highly unusual occurrence of three cases within 22 days in the same institute.

Analysis of TCR sequencing reveals expansion of similar clones across multiple samples. The similarity between TCR found in the PM cardiac muscle, skeletal muscle and tumour of patient 3 and the skeletal muscle of patient 1 is significantly higher than when compared to the TCR repertoires sequenced from muscle biopsies of IIM or than seen between these tissues and resected melanomas from pre-2020. As such, this suggests clonal expansion of a subset of public clones not found in other forms of myositis, indicating possible recognition of a distinct set of antigens common to both muscle and tumour. Some of these TCR are known to be Spike-reactive, and muscle-infiltrating TCR are enriched for this epitope compared to the peripheral blood of the same patient.

Myocarditis following ICB therapy is well-described^5,21,22^, as is an association with mRNA COVID-19 vaccines^9–12^. Cardiomyocytes express high levels of PD-L1 which is upregulated in the context of myocardial injury, serving to abrogate severe myocarditis^23–27^. Similarly, PDL-1 expression in the inflamed tumor microenvironment curtails anti-tumor T cell activity. Expression of mRNA vaccine derived antigen in muscle triggers local inflammation which may elicit a degree of muscle autoimmunity. In the context of subsequent early anti-PD1 treatment, the physiological feedback through PD-L1:PD-1 ligation and peripheral tolerance mechanisms preventing development of systemic autoimmunity may be overcome. We postulate these cases may represent the consequence of *de novo* anti-PD1 infusion post-boost vaccination, revealing anti-muscle autoimmunity with concomitant myaesthenia gravis symptoms. An increase in incidence of myocarditis post-ICB has been noted by others since the beginning of the COVID-19 pandemic^28^ and it is possible similar cases with this unusual clinical presentation have been overlooked.

Notably, these data are observational and causality is not assigned. Moreover, the TCRseq data is based on CDR3B amino acid sequence rather than the complete chain. We suspect the high number of putative Spike-recognising TCR reflects, at least in-part, extensive prior analysis of this epitope and disproportionate representation in databases. In keeping with this, there was a generalised enrichment of TCR recognising anti-viral epitopes. Finally, we have recently described the association of the minor allele of rs16906115, intragenic to *IL7*, with the development of IRAEs to ICB^29,30^. We tested the three index cases for carriage of this allele, but found all to be homozygous for the major allele, arguing against common genetic predisposition and again indicating a recent shared environmental factor.

Any potential clinical risk identified by this study needs to be considered in the context of the significant benefits COVID-19 vaccination provides, especially in patients with cancer. In addition to the temporal clustering of these cases over a 2 month period, these cases are all characterised by the infusion of a 42 day (high) dose of Pembrolizumab in a 12-week window post-booster vaccination with toxicities developing prior to any subsequent infusions. We would urge physicians to be aware of this association, with a low threshold for assessment and monitoring of ECG changes, blood enzymes and clinical status. Consideration should be given to initiating three-weekly Pembrolizumab prior to switching after four cycles and booster vaccination being given post-initiation of ICB treatment as opposed to in the prior weeks.

## Data Availability

All data produced in the present study are available upon reasonable request to the authors

## Acknowledgements

We are very grateful to all patients and their families who contributed samples and participated in the study. We thank all the staff of the Day Treatment Unit, Oxford Cancer Centre, and The Brodey Centre at the Horton General Hospital. We are grateful to all the staff of the Oxford University Hospitals NHS Foundation Trust, as well as the staff of the Oxford Radcliffe Biobank (ORB) and Churchill Hospital Sample Handling Lab.

## Supplemental information

### Supplemental Tables

**Table S1.** Samples on which TCRseq was performed. PBMC - peripheral blood mononuclear cell; PM - post mortem; IMNM - immune-mediated necrotising myositis; DM - dermatomyositis; IBM - inclusion body myositis; AyS - anti synthetase syndrome; IIM - idiopathic inflammatory myositis. Myositis control samples came from Montagne *et al*^18^

| Reported (index) cases |  | Myositis controls (Montagne) -<br>Fresh Frozen Muscle Biopsies |
| --- | --- | --- |
| Patient 1 | PBMCs (post treatment) | IMNM |
|  | Fresh muscle biopsy (post treatment) | DM |
| Patient 2 | PBMCs (post treatment) | IBM |
| Patient 3 | PBMCs (pre treatment) | ASyS |
|  | PM heart (post treatment) | Non-IIM |
|  | PM muscle (post treatment) | Healthy |
|  | PM tumour (post treatment) |  |

**Table S2.** Resection date and pathological detail for 12 resected melanoma specimens. Slides were microscopically examined to identify areas of melanoma before macrodissection and deparafinisation. TCR sequencing was then performed as described in Methods.

| Tumour block | Specimen resection date | Specimen detail |
| --- | --- | --- |
| T51 | 19/01/2017 x2 | Excised melanoma from skin (x2 lesions) |
| T61 | 22/02/2017 | Excised melanoma from skin |
| T64 | 07/04/2017 | Melanoma deposit within lymph node |
| T90 | 10/03/2017 | Melanoma deposit within lymph node |
| T101 | 19/02/2018 | Excised melanoma from skin |
| T104 | 23/02/2017, 12/05/2017, 04/01/2018 | Excised melanoma from skin (x1 lesion) and melanoma deposit within lymph node (x2 lesions) |
| T108 | 27/06/2017 | Excised melanoma from skin |
| T132 | 07/08/2018 | Excised invasive melanoma from muscle |

Table S3. THIS IS A CSV FILE. Chain information for patient 1 PBMC and muscle biopsy with IEDB organism, epitope and score.

### Supplemental Figures

**Figure S1.**
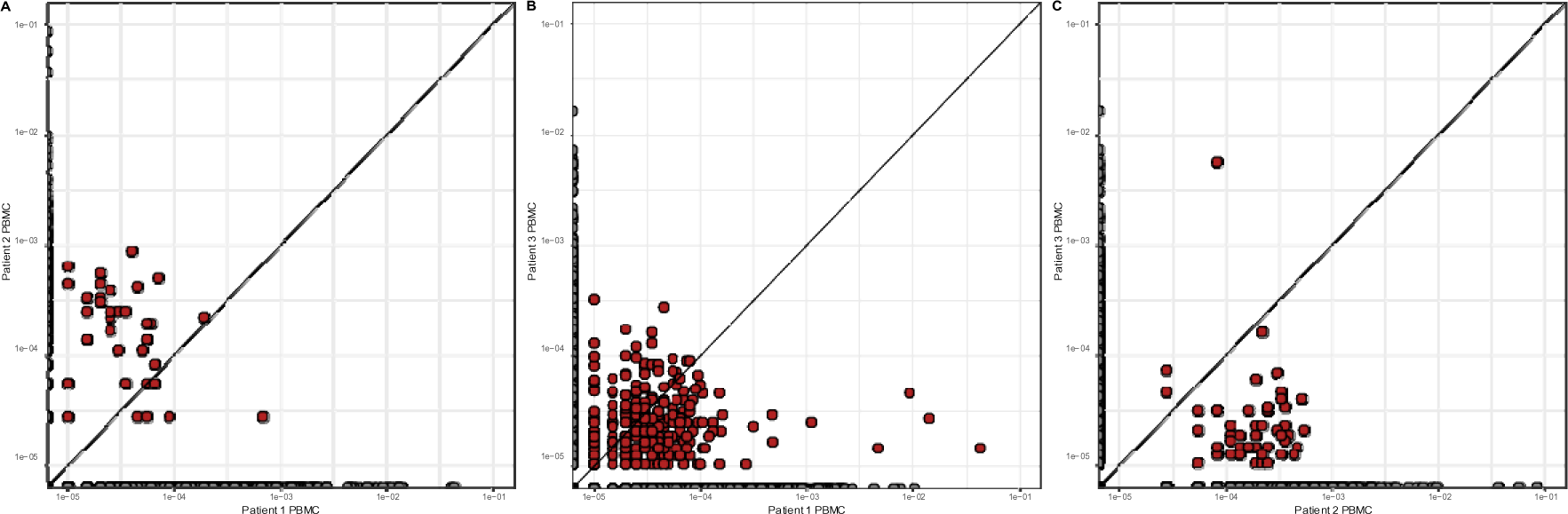
(A-C) Dot plot showing proportion of repertoire occupied by T cell clones in peripheral blood samples (as labelled).

**Figure S2.**
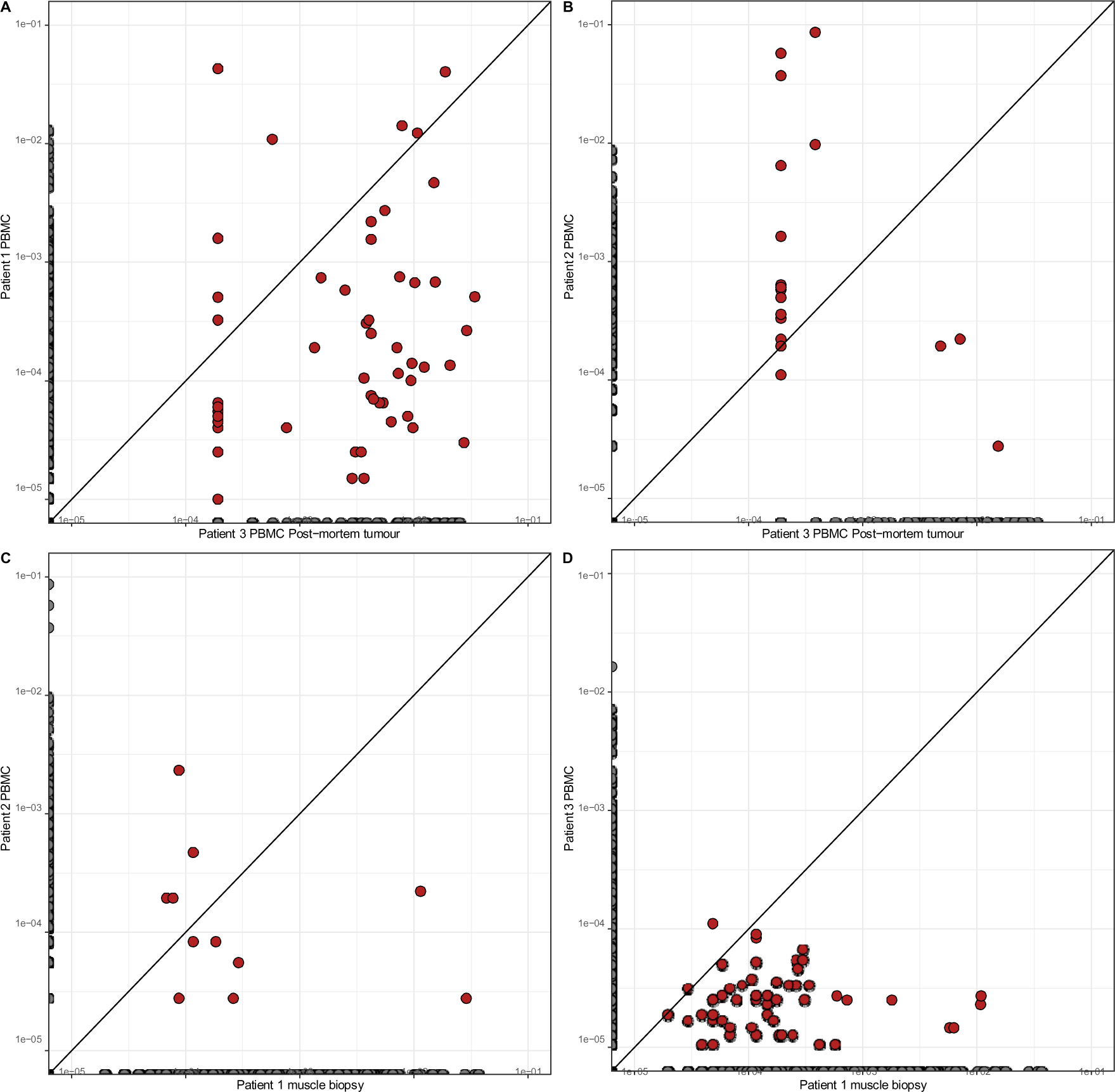
(A-B) Dot plot showing proportion of repertoire occupied by T cell clones in PM tumour sample from patient 3 compared to peripheral blood of patients 1 and 2 (as labelled). (C-D) as for (A-B) but comparing fresh muscle biopsy from patient 1 to peripheral blood of patients 2 and 3 (as labelled).

**Figure S3.**
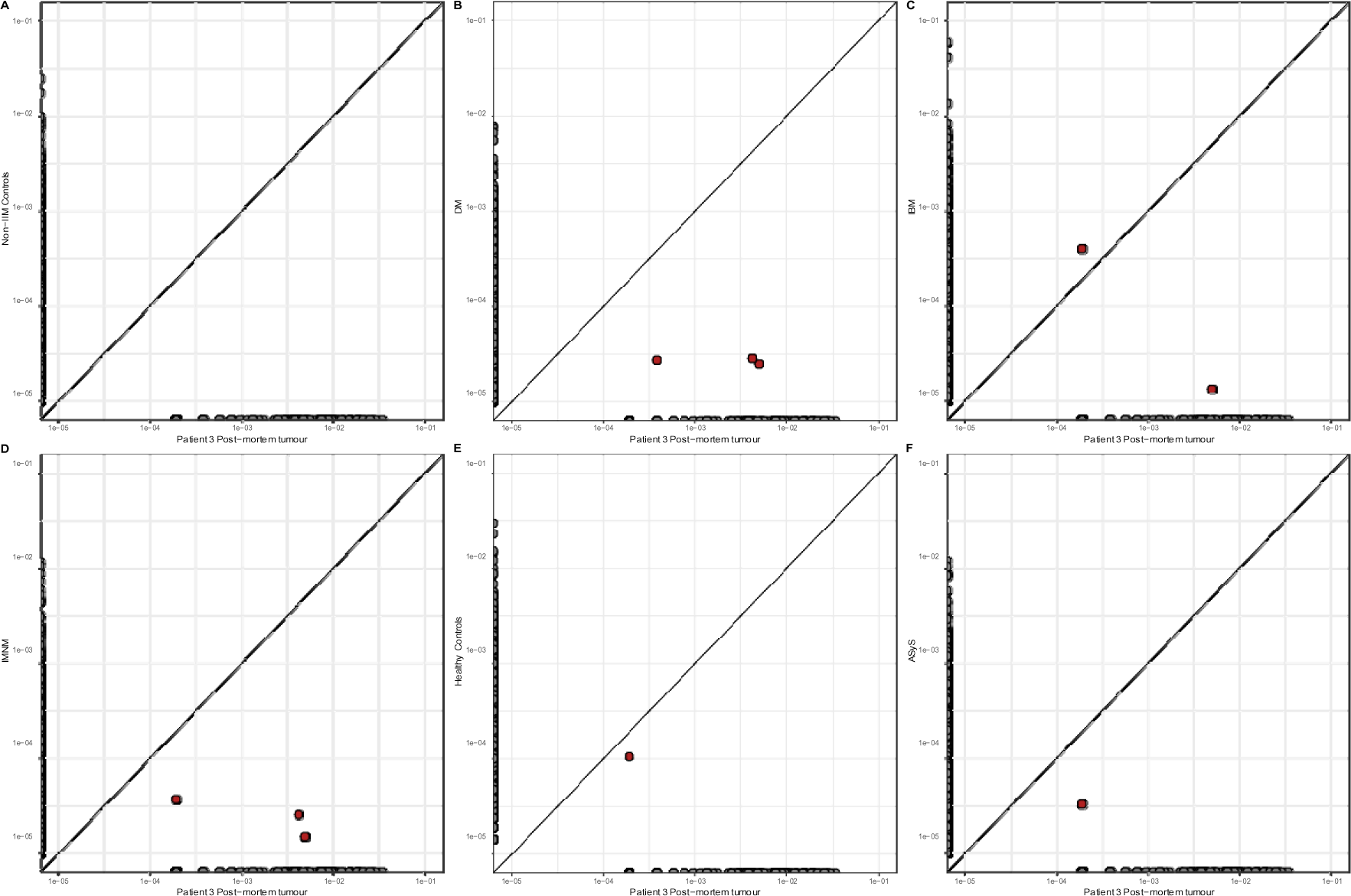
(A-F) Dot plot showing proportion of repertoire occupied by T cell clones in PM tumour sample from patient 3 compared to those found in muscle biopsies taken from patients with IIM^18^. N.B. patients with IIM are grouped by condition, each condition containing multiple individuals. IMNM - immune-mediated necrotising myositis; DM - dermatomyositis; IBM - inclusion body myositis; AyS - anti synthetase syndrome; IIM - idiopathic inflammatory myositis.

**Figure S4.**
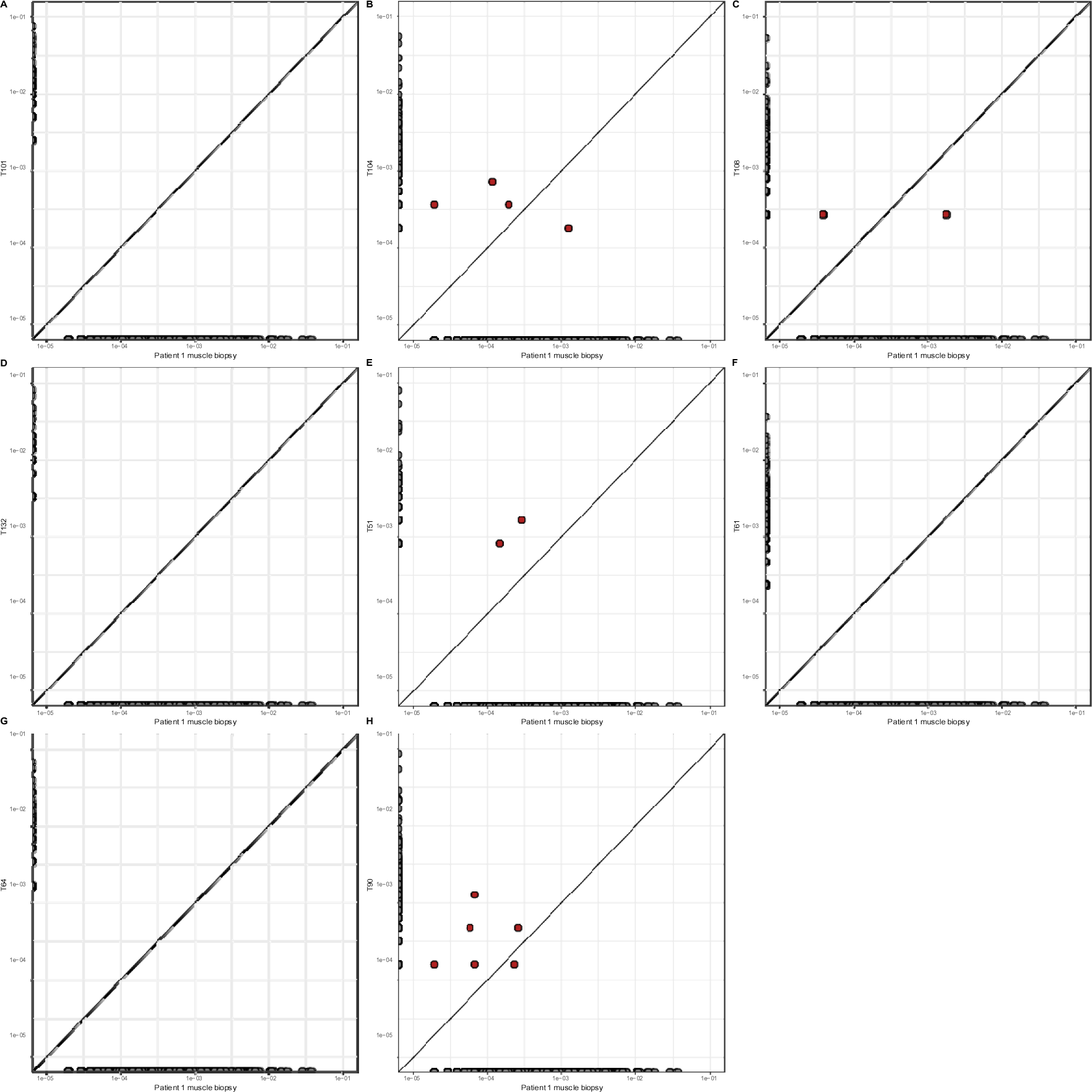
(A-H) Dot plot showing proportion of repertoire occupied by T cell clones in fresh muscle biopsy from patient 1 compared to those found in resected melanomas from before 2020.

**Figure S5.**
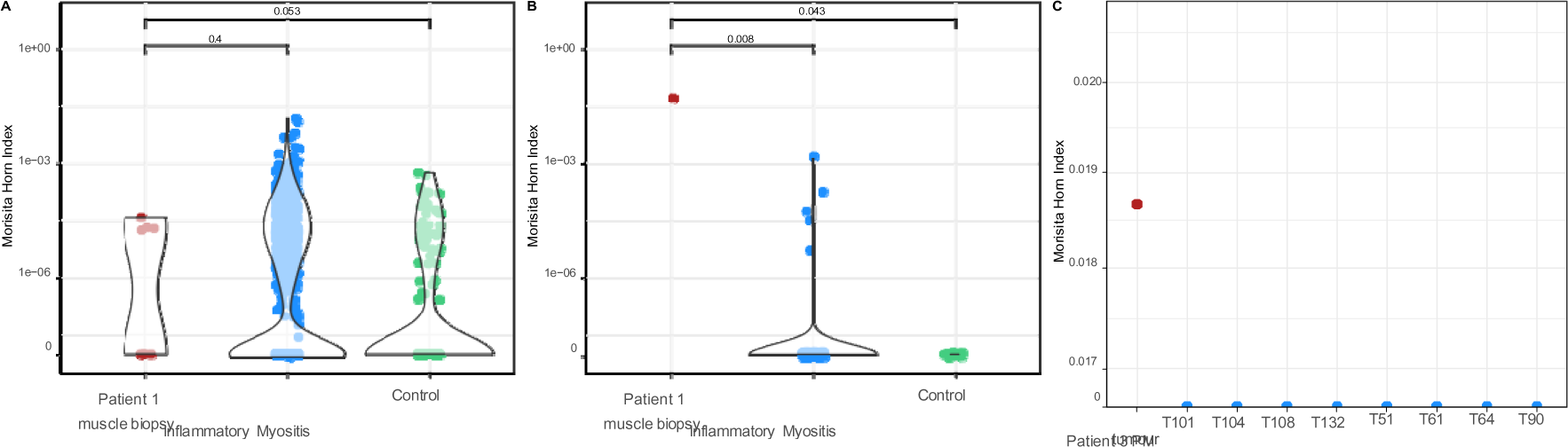
(A) Morisita-Horn (MH) index for the TCR repertoire overlap between resected melanomas from eight patients from the original cohort (table S2) and skeletal muscle biopsies as labelled (N.B. repertoires found in inflammatory myositis biopsies and control biopsies came from Montagne *et al*^18^). (B) as for (A) but MH index for TCR repertoire overlap between PM cardiac muscle from patient 3 and muscle biopsies as labelled. (C) MH index showing TCR repertoire overlap between the PM cardiac muscle from patient 3, the PM tumour from patient 3 and resected melanomas from eight patients pre-2020 (table S2).

